# Cumulative Remnant Cholesterol as a causal risk factor for Ischemic Heart Disease: A Prospective Cohort Study

**DOI:** 10.1101/2023.10.26.23297647

**Authors:** Xuemei Zhao, Yifei Wang, Wei Li, Haibo Gao, Haotian Wu, Jiaxin Yu, Guiping Wang, Shouling Wu, Xiaokun Liu

**Affiliations:** Department of Cardiology, Tangshan Workers’ Hospital,China; Hebei Medical University, China; North China University of Science and Technology, Tangshan, China; Department of Cardiology, Kailuan Hospital, North China University of Science and Technology, Tangshan City, China

**Keywords:** Remnant cholesterol, Ischemic heart disease, Cumulative exposure, Cohort study, Prevention

## Abstract

**Background:** While previous studies have established a significant correlation between baseline remnant cholesterol (RC) and ischemic heart disease (IHD), the enduring impact of RC on incident IHD remains to be elucidated. This study aimed to investigate the association between cumulative remnant cholesterol(cumRC) and IHD susceptibility.

**Methods:** Participating from the Kailuan Study (2006-2010) were enrolled, excluding those with prior myocardial infarction, coronary artery revascularization and cancer across three consecutive examinations. The cumRC derived by multiplying the average RC with the interval between the two consecutive assessments. Participants were segmented into quartiles based on cumRC levels: Q1 (cumRC<2.69mmol/l); Q2 (2.69≤cumRC<4.04mmol/l); Q3(4.04≤cumRC<5.65mmol/l) and Q4 (cumRC≥5.65mmol/l). The correlation between cumRC and IHD risk was ascertained by using multivariable Cox proportional hazard models.

**Result:** The analysis encompassed 42,639 participants. Over an average tracking period of 9.97 years, 1,205 instances of IHD were identified. IHD susceptibility augmented with rising cumRC quartiles. After adjusting for potential confounders, the hazard ratios for IHD events were 1.06 (0.88-1.29) for Q2, 1.30 (1.08-1.56) for Q3 and 1.69 (1.42-2.01) for Q4, relative to Q1. Elevated cumRC was significantly associated with a heightened IHD risk, a trend consistent in both subgroup and sensitivity analyses.

**Conclusion:** Elevated cumRC significantly correlates with a higher risk of IHD, suggesting that consistent monitoring and regulation of RC might be instrumental in IHD prevention.

## INTRODUCTION

Low-density lipoprotein cholesterol (LDL-C) is an independent risk factor for coronary heart disease.[1, 2] A confluence of epidemiological, genetic, and clinical intervention research underscores that elevated LDL-C concentrations precipitate atherosclerotic cardiovascular incidents (ASCVD).[3] Prior research posits that a reduction of 1 mmol/L in LDL-C corresponds to a 25% diminution in major cardiovascular occurrences.[4] Yet, certain investigations highlight that even with diminished LDL-C concentrations, there persists a tangible cardiovascular disease risk, potentially attributable to remnant cholesterol (RC).[5-7] A Mendelian randomization study discerned a staggering 2.8-fold surge in ischemic heart disease (IHD) risk with every 1 mmol/l augmentation in RC, suggesting that escalated RC concentrations are instrumental in IHD onset.[8] Furthermore, a plethora of research consistently indicates that heightened RC levels correlate with an amplified risk of cardiovascular diseases.[6, 9-12] A 2021 cohort analysis revealed that within the broader populace, RC concentrations were intrinsically linked to ASCVD, independent of the conventional cardiovascular risk markers like LDL-C, non-HDL-C, or apoB concentrations.[13].

While prior research has delved into the correlation between RC levels and IHD, the emphasis has predominantly been on singular RC measurements. Such isolated measurements can be skewed by a myriad of variables, including age, lifestyle, and medicinal interventions, rendering them inadequate in capturing the enduring influence of RC levels on IHD. Several studies have adopted a more comprehensive approach, gauging the nexus between diseases and risk factors via recurrent long-term risk factor assessments. For instance, cumulative hyperlipidemia exposure during adolescence has been shown to proportionally escalate the likelihood of subsequent cardiovascular incidents.[14] The 2020 CARDIA study discerned that cardiovascular event risks are contingent upon cumulative LDL-C exposure.[15] Following this, a 2021 prospective cohort study ascertained that elevated cumulative LDL-C exposure during one’s youth and midlife phases correlates with a heightened propensity for coronary heart disease onset.[16] Yet, both national and international research spheres exhibit a conspicuous void in studies employing repeated RC level measurements to elucidate the long-term RC exposure’s relationship with IHD. The Kailuan study stands out, having meticulously tracked and amassed extended multi-lipid data from participants through recurrent long-term follow-ups, thereby securing multiple RC level readings over time. Consequently, this research leveraged a prospective cohort methodology to probe the ramifications of cumulative remnant cholesterol (cumRC) exposure on IHD within the Kailuan Study cohort.

## METHODS

### Study participants

Participants for this investigation were sourced from the Kailuan Study cohort. The Kailuan Study (accession number: ChiCTR-TNC- 11001489) represents a comprehensive prospective cohort analysis focusing on both active and retired personnel of the Kailuan Group. Detailed study methodologies and procedures are elaborated in prior publications by the team.[17, 18] Commencing in 2006, hospitals affiliated with the Kailuan Group initiated routine health examinations and questionnaire-based surveys, with biennial follow-up sessions. For this study, we considered 57,927 employees who consistently partook in the health evaluations orchestrated by the Kailuan Group in 2006, 2008, and 2010. Out of these, 44,520 individuals with a comprehensive lipid profile were incorporated into the research. Exclusions were made for participants presenting with myocardial infarction and coronary artery revascularisation during the 2006, 2008, and 2010 health assessments (n=1524), as well as those with a prior tumour diagnosis (n=357). Consequently, a cohort of 42,639 participants was finalized for this study.

This research adhered to the guidelines stipulated by the Declaration of Helsinki and received approval from the Ethics Committee of Kailuan General Hospital (200608). All participants voluntarily joined the study and furnished their written informed consent.

### Data Collection and definitions

Participant details were gleaned via a questionnaire, capturing demographics such as age, gender, ethnicity, marital status, and educational background. It also recorded lifestyle habits like smoking, alcohol consumption, and physical activity, along with medical history and medication details. The Body Mass Index (BMI) was computed by dividing weight (in kg) by the square of height (in m^2).

Blood pressure from the right brachial artery was gauged using a precision mercury sphygmomanometer. Prior to this, participants refrained from smoking or consuming caffeine for 30 minutes and rested in a seated position for 15 minutes. Three successive readings were taken, spaced 1-2 minutes apart, with the average value recorded.

A 5ml fasting venous blood sample (post an 8-hour fast) was drawn from the brachial vein. Post-centrifugation, the serum’s upper layer was isolated. Biochemical markers, including Fasting Blood Glucose (FBG), Triglycerides (TG), Total Cholesterol (TC), High-Density Lipoprotein Cholesterol (HDL-C), and Low-Density Lipoprotein Cholesterol (LDL-C), were analyzed using a Hitachi 7600 automatic biochemical analyzer at Kailuan General Hospital’s Central Laboratory.

Individuals who had smoked for over a year, averaging at least one cigarette daily and had smoked in the preceding year, were classified as ’ever-smokers’. ’Ever-drinkers’ were those who had consumed alcohol for over a year, averaging at least 100 ml/day of 50% alcohol volume liquor, and had consumed alcohol in the past year. Hypertension was characterized by a systolic blood pressure of ≥140 mmHg and/or a diastolic blood pressure of ≥90 mmHg, or those on antihypertensive medication despite having lower readings.[19] Diabetes mellitus was identified by a fasting blood glucose level of ≥7.0 or those with a confirmed diabetes diagnosis or on glucose-lowering medication, even if their fasting blood glucose was <7.0 mmol/L.[20]

### RC and cumRC

The concentration of Remnant Cholesterol (RC) was determined using VerboA’s proposed formula[8]: RC = TC - HDL-C - LDL-C. The cumulative RC (cumRC) was computed as follows: cumRC = [(RC2006 + RC2008) / 2 × time1-2] + [(RC2008 + RC2010) / 2 × time2-3]. Here, RC2006, RC2008, and RC2010 represent the RC values from the initial, baseline, and third evaluations, respectively. The terms time1-2 and time2-3 denote the specific intervals, in years, between successive visits for each participant.[21] Based on the cumRC quartiles, participants were segmented into four groups: Q1 (cumRC < 2.69mmol/l), Q2 (2.69 ≤ cumRC < 4.04mmol/l), Q3 (4.04 ≤ cumRC < 5.65mmol/l), and Q4 (cumRC ≥ 5.65mmol/l).

### Follow-up and outcomes

From the onset of participant enrollment in 2006, we conducted annual follow-ups to collect endpoint event data. Disease classifications and diagnostic details were sourced from participants’ electronic medical records during hospital visits, referencing the International Classification of Diseases and Related Health Problems, Tenth Revision (ICD-10). Mortality data were acquired from provincial vital statistics offices. In this research, the primary outcome was identified as ischemic heart disease, which includes myocardial infarction and coronary artery revascularization. The diagnosis of myocardial infarction was ascertained through clinical symptoms, cardiac enzyme metrics, and evolving electrocardiogram patterns[22]. For participants with multiple endpoint events, the earliest event determined the outcome. For those without ischemic heart disease incidents, the final follow-up was marked as December 31, 2021.

### Statistical analysis of data

All analyses were executed using SAS version 9.4 (SAS Institute, Cary, North Carolina). Continuous data with normal distribution are denoted as mean ± standard deviation, and intergroup variations were assessed via one-way analysis of variance. Non-normally distributed continuous data are represented as median (P25, P75), with intergroup differences evaluated using the Kruskal-Wallis non-parametric test. Categorical data are depicted as frequencies (percentages), and group disparities were determined using the χ2 test.

The Kaplan-Meier approach estimated the cumulative incidence rates of newly diagnosed IHD during the follow-up for each cumRC group, with group comparisons made using the log-rank test. Multivariable Cox proportional hazards regression analysis, with IHD as the dependent variable and cumRC as the independent variable, gauged the influence of distinct cumRC quartiles on IHD, referencing the Q1 quartile. Model 1 adjusted for age and gender; Model 2 incorporated additional factors; Model 3 further adjusted for LDL-C and HDL-C; Model 4 considered the initial RC measurement; Model 5 included the baseline RC level. To account for the potential influence of lipid-lowering medications on RC levels, Model 6 expanded on Model 3 by adjusting for the introduction of new lipid-lowering medications during follow-up. An analysis evaluated the effect of cumRC on IHD for each one standard deviation (SD) increase. To ensure result robustness, two sensitivity analyses were conducted: 1) Excluding events from the initial 2 years of follow-up to mitigate potential reverse causation, and 2) Excluding participants on medication and reanalyzing. Stratified analyses based on gender, smoking status, and diabetes were also conducted. Given the competing risk of non-IHD death, a competing risk survival model was employed. The C-index, IDI, and NRI compared the predictive values of RC2006, RC2010, and cumRC for IHD risk. All tests were 2-sided, with P<0.05 denoting statistical significance.

## RESULT

### Comparison of Baseline Characteristics of Study Participants

In this study, 42,639 participants were deemed eligible, with an average age of 53.46±11.86 years. The cohort comprised 32,507 males (76.43%) and 10,027 females (23.57%). Participants were evenly distributed across the cumRC quartiles: Q1 group had 10,659 participants (25%), Q2 group had 10,660 (25%), Q3 group had 10,660 (25%), and Q4 group also had 10,660 (25%). A notable observation was that individuals with elevated cumRC levels displayed higher metrics for systolic blood pressure, triglycerides, total cholesterol, and fasting blood glucose than their counterparts with lower cumRC. Furthermore, a larger percentage of those in the higher cumRC bracket were on antihypertensive and lipid-lowering medications. They also had a higher body mass index (BMI) (all P < 0.05). These findings are summarized in (Table 1).

**Table 1.**
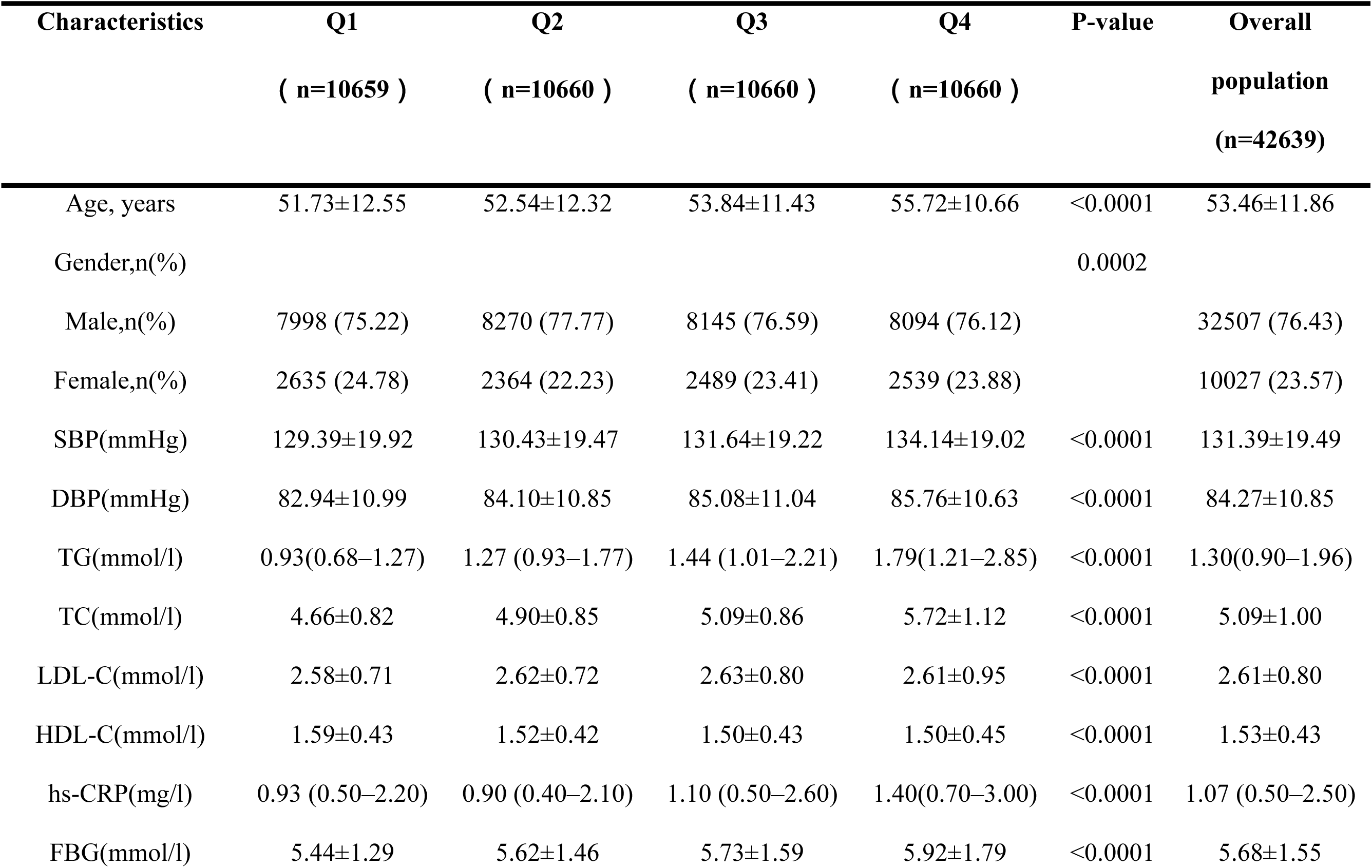
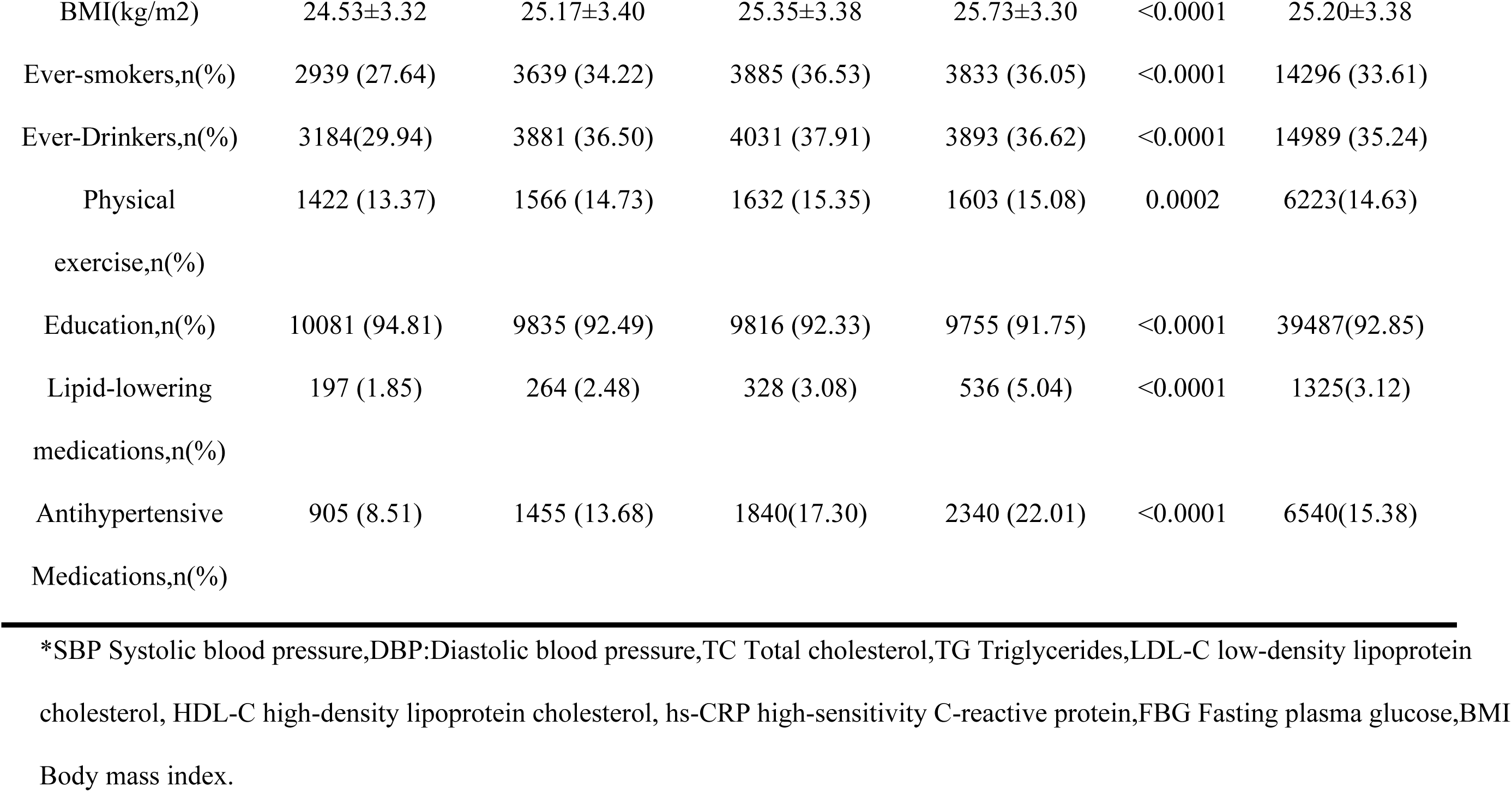
Baseline characteristics of 42639populations acrroding to 4 groups divided by cumRC quartile.

| Characteristics | Q1<br>( n=10659 ) | Q2<br>( n=10660 ) | Q3<br>( n=10660 ) | Q4<br>( n=10660 ) | P-value | Overall<br>population<br>(n=42639) |
| --- | --- | --- | --- | --- | --- | --- |
| Age, years | 51.73±12.55 | 52.54±12.32 | 53.84±11.43 | 55.72±10.66 | <0.0001 | 53.46±11.86 |
| Gender,n(%) |  |  |  |  | 0.0002 |  |
| Male,n(%) | 7998 (75.22) | 8270 (77.77) | 8145 (76.59) | 8094 (76.12) |  | 32507 (76.43) |
| Female,n(%) | 2635 (24.78) | 2364 (22.23) | 2489 (23.41) | 2539 (23.88) |  | 10027 (23.57) |
| SBP(mmHg) | 129.39±19.92 | 130.43±19.47 | 131.64±19.22 | 134.14±19.02 | <0.0001 | 131.39±19.49 |
| DBP(mmHg) | 82.94±10.99 | 84.10±10.85 | 85.08±11.04 | 85.76±10.63 | <0.0001 | 84.27±10.85 |
| TG(mmol/l) | 0.93(0.68–1.27) | 1.27 (0.93–1.77) | 1.44 (1.01–2.21) | 1.79(1.21–2.85) | <0.0001 | 1.30(0.90–1.96) |
| TC(mmol/l) | 4.66±0.82 | 4.90±0.85 | 5.09±0.86 | 5.72±1.12 | <0.0001 | 5.09±1.00 |
| LDL-C(mmol/l) | 2.58±0.71 | 2.62±0.72 | 2.63±0.80 | 2.61±0.95 | <0.0001 | 2.61±0.80 |
| HDL-C(mmol/l) | 1.59±0.43 | 1.52±0.42 | 1.50±0.43 | 1.50±0.45 | <0.0001 | 1.53±0.43 |
| hs-CRP(mg/l) | 0.93 (0.50–2.20) | 0.90 (0.40–2.10) | 1.10 (0.50–2.60) | 1.40(0.70–3.00) | <0.0001 | 1.07 (0.50–2.50) |
| FBG(mmol/l) | 5.44±1.29 | 5.62±1.46 | 5.73±1.59 | 5.92±1.79 | <0.0001 | 5.68±1.55 |
| BMI(kg/m<sup>2</sup>) | 24.53±3.32 | 25.17±3.40 | 25.35±3.38 | 25.73±3.30 | <0.0001 | 25.20±3.38 |
| Ever-smokers,n(%) | 2939 (27.64) | 3639 (34.22) | 3885 (36.53) | 3833 (36.05) | <0.0001 | 14296 (33.61) |
| Ever-Drinkers,n(%) | 3184(29.94) | 3881 (36.50) | 4031 (37.91) | 3893 (36.62) | <0.0001 | 14989 (35.24) |
| Physical<br>exercise,n(%) | 1422 (13.37) | 1566 (14.73) | 1632 (15.35) | 1603 (15.08) | 0.0002 | 6223(14.63) |
| Education,n(%) | 10081 (94.81) | 9835 (92.49) | 9816 (92.33) | 9755 (91.75) | <0.0001 | 39487(92.85) |
| Lipid-lowering<br>medications,n(%) | 197 (1.85) | 264 (2.48) | 328 (3.08) | 536 (5.04) | <0.0001 | 1325(3.12) |
| Antihypertensive<br>Medications,n(%) | 905 (8.51) | 1455 (13.68) | 1840(17.30) | 2340 (22.01) | <0.0001 | 6540(15.38) |

\*SBP Systolic blood pressure,DBP:Diastolic blood pressure,TC Total cholesterol,TG Triglycerides,LDL-C low-density lipoprotein cholesterol, HDL-C high-density lipoprotein cholesterol, hs-CRP high-sensitivity C-reactive protein,FBG Fasting plasma glucose,BMI Body mass index.

### The relationship between incident IHD and cumRC

Over a median follow-up duration of 9.97 years, 1205 participants (2.82%) developed IHD. This included 499 cases of myocardial infarction and 706 instances of coronary artery revascularization. The Kaplan-Meier curves revealed a stepwise increase in the cumulative incidence of IHD as we move from the Q1 to Q4 groups (Figure 1a, log-rank test, p < 0.0001). This trend was consistent for both myocardial infarction and coronary artery revascularization (Figure 1b and 1c respectively, log-rank test, p < 0.0001). After accounting for potential confounders, cumRC emerged as a significant predictor for IHD, independent of LDL-C and HDL-C levels. When using the Q1 group as a baseline, the risk of IHD escalated with rising cumRC (P for trend < 0.0001). The Q4 group exhibited the highest risk (HR, 1.69; 95% CI, 1.42-2.01). On further stratifying IHD into myocardial infarction and coronary artery revascularization, the findings remained consistent. Moreover, with every 1-SD (2.34 mmol/l) increase in cumRC, there was a 19% surge in IHD risk compared to the Q1 reference group (P<0.0001). These results are detailed in Table 2.

**Figure 1.**
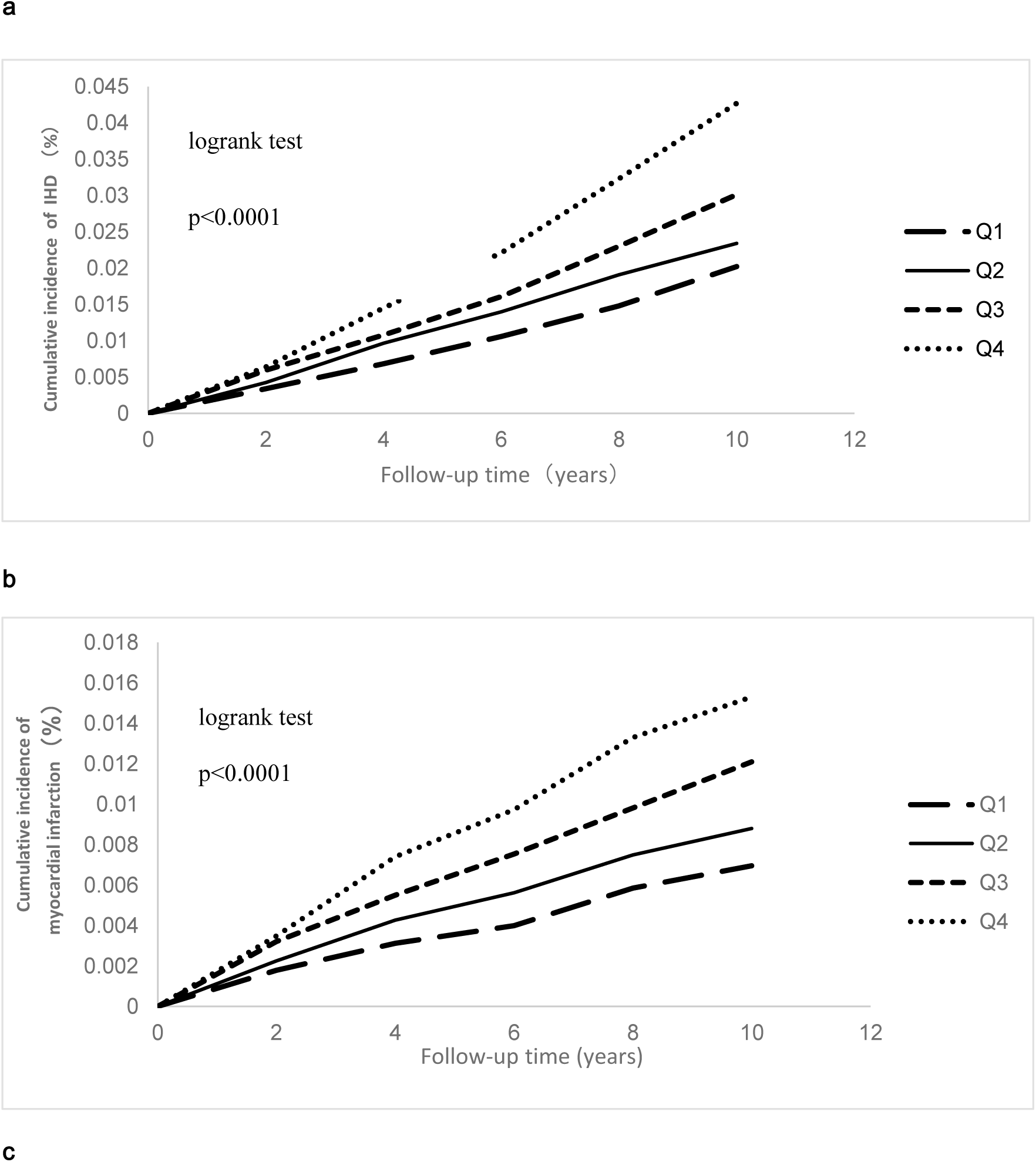
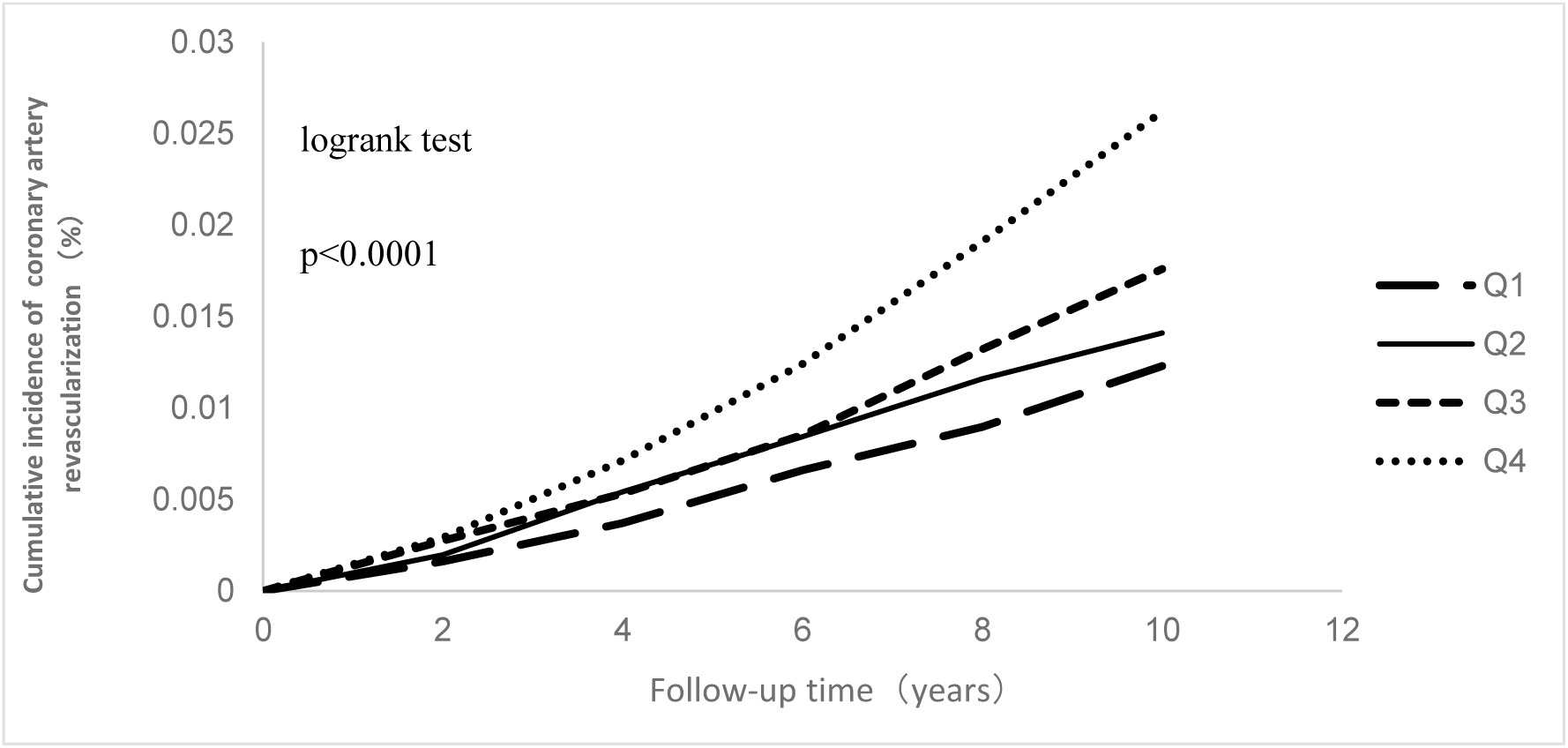
Cumulative incidence curve of Ischemic heart disease in four groups

**Table 2.** COX proportional hazard model analysis of different cumRC groups and the incidence Ischemic heart disease.

|  | **Q1**<br>**(n=10659)** | **Q2**<br>**(n=10660)** | **Q3**<br>**(n=10660)** | **Q4**<br>**(n=10660)** | **P for trend** | **SD** | **P value for SD** |
| --- | --- | --- | --- | --- | --- | --- | --- |
| **Ischemic heart disease** |  |  |  |  |  |  |  |
| No of case. | 205 | 243 | 315 | 442 |  |  |  |
| IR(/Per 1000 Person Years) | 2.00 | 2.11 | 2.78 | 4.04 |  |  |  |
| Model1HR(95%CI) | 1 [Reference] | 1.16(0.97-1.40) | 1.48(1.24-1.76) | 2.00(1.70-2.37) | <0.0001 | 1.24(1.19-1.30) | <0.0001 |
| Model2HR(95%CI) | 1 [Reference] | 1.09(0.90-1.31) | 1.32(1.10-1.58) | 1.68(1.42-2.00) | <0.0001 | 1.19(1.13-1.25) | <0.0001 |
| Model3HR(95%CI) | 1 [Reference] | 1.06(0.88-1.29) | 1.30(1.08-1.56) | 1.69(1.42-2.01) | <0.0001 | 1.19(1.14-1.25) | <0.0001 |
| Model4HR(95%CI) | 1 [Reference] | 1.03(0.85-1.25) | 1.21(1.00-1.47) | 1.49(1.20-1.84) | <0.0001 | 1.14(1.07-1.22) | <0.0001 |
| Model5HR(95%CI) | 1 [Reference] | 1.03(0.85-1.24) | 1.22(1.02-1.47) | 1.42(1.18-1.72) | <0.0001 | 1.11(1.05-1.18) | 0.0006 |
| Model6HR(95%CI) | 1 [Reference] | 1.08(0.89-1.30) | 1.24(1.03-1.49) | 1.39(1.17-1.65) | <0.0001 | 1.12(1.07-1.18) | <0.0001 |
| **Myocardial infarction** |  |  |  |  |  |  |  |
| No of case. | 81 | 99 | 138 | 181 |  |  |  |
| IR(/Per 1000 Person Years) | 0.72 | 0.88 | 1.23 | 1.64 |  |  |  |
| Model1HR(95%CI) | 1 [Reference] | 1.19(0.89-1.60) | 1.64(1.25-2.16) | 2.04(1.57-2.65) | <0.0001 | 1.23(1.15-1.31) | <0.0001 |
| Model2HR(95%CI) | 1 [Reference] | 1.17(0.87-1.59) | 1.57(1.18-2.09) | 1.89(1.44-2.48) | <0.0001 | 1.20(1.11-1.29) | <0.0001 |
| Model3HR(95%CI) | 1 [Reference] | 1.13(0.84-1.53) | 1.53(1.15-2.03) | 1.88(1.43-2.46) | <0.0001 | 1.19(1.11-1.28) | <0.0001 |
| Model4HR(95%CI) | 1 [Reference] | 1.11(0.81-1.50) | 1.47(1.08-1.99) | 1.74(1.24-2.44) | 0.0003 | 1.15(1.05-1.27) | 0.0040 |
| Model5HR(95%CI) | 1 [Reference] | 1.09(0.80-1.47) | 1.43(1.07-1.91) | 1.55(1.15-2.09) | 0.0007 | 1.11(1.01-1.21) | 0.0379 |
| Model6HR(95%CI) | 1 [Reference] | 1.14(0.83-1.55) | 1.52(1.14-2.02) | 1.66(1.26-2.20) | <0.0001 | 1.16(1.07-1.25) | 0.0002 |
| **Coronary artery revascularization** |  |  |  |  |  |  |  |
| No of case. | 124 | 144 | 177 | 261 |  |  |  |
| IR(/Per 1000 Person Years) | 1.21 | 1.41 | 1.74 | 2.61 |  |  |  |
| Model1HR(95%CI) | 1 [Reference] | 1.14(0.90-1.45) | 1.38(1.10-1.73) | 1.97(1.59-2.44) | <0.0001 | 1.25(1.19-1.32) | <0.0001 |
| Model2HR(95%CI) | 1 [Reference] | 1.03(0.80-1.31) | 1.17(0.92-1.48) | 1.55(1.24-2.94) | <0.0001 | 1.18(1.11-1.26) | <0.0001 |
| Model3HR(95%CI) | 1 [Reference] | 1.02(0.80-1.30) | 1.16(0.92-1.47) | 1.56(1.25-1.95) | <0.0001 | 1.18(1.11-1.26) | <0.0001 |
| Model4HR(95%CI) | 1 [Reference] | 0.97(0.75-1.24) | 1.06(0.82-1.36) | 1.32(1.00-1.74) | 0.0258 | 1.13(1.04-1.23) | 0.0045 |
| Model5HR(95%CI) | 1 [Reference] | 0.99(0.77-1.26) | 1.10(0.87-1.40) | 1.34(1.05-1.72) | 0.0077 | 1.12(1.03-1.21) | 0.0060 |
| Model6HR(95%CI) | 1 [Reference] | 1.00(0.78-1.28) | 1.04(0.82-1.32) | 1.18(0.94-1.47) | 0.1068 | 1.08(1.01-1.16) | 0.0224 |

SD,standard deviation.CI, confidence interval; HR ,hazard ratio.Model 1 ,adjusted for age and gender.Model 2, adjusted for variables in model 1 plus smoking,drinking,physical activity, level of education,SBP,FBG,BMI,lipid-lowering drugs, antihypertensive drugs,hs-CRP.Model 3,adjusted for variables in model 2 plus HDL-C, LDL-C.Model 4,adjusted for variables in model 3 plus the initial RC measurement.Model 5,adjusted for variables in model 3 plus the baseline RC level.Model 6,adjusted for variables in model 3 plus the introduction of new lipid-lowering medications during follow-up.

### Sensitivity Analysis and Competing Risk of Death

The outcomes remained consistent when we excluded cases of IHD that occurred within the first 2 years of follow-up or those that received medical treatment (Table 3). Additionally, when employing the Fine-Gray model to evaluate the relationship between cumRC and IHD, the findings remained stable (Table 4).

**Table 3.**
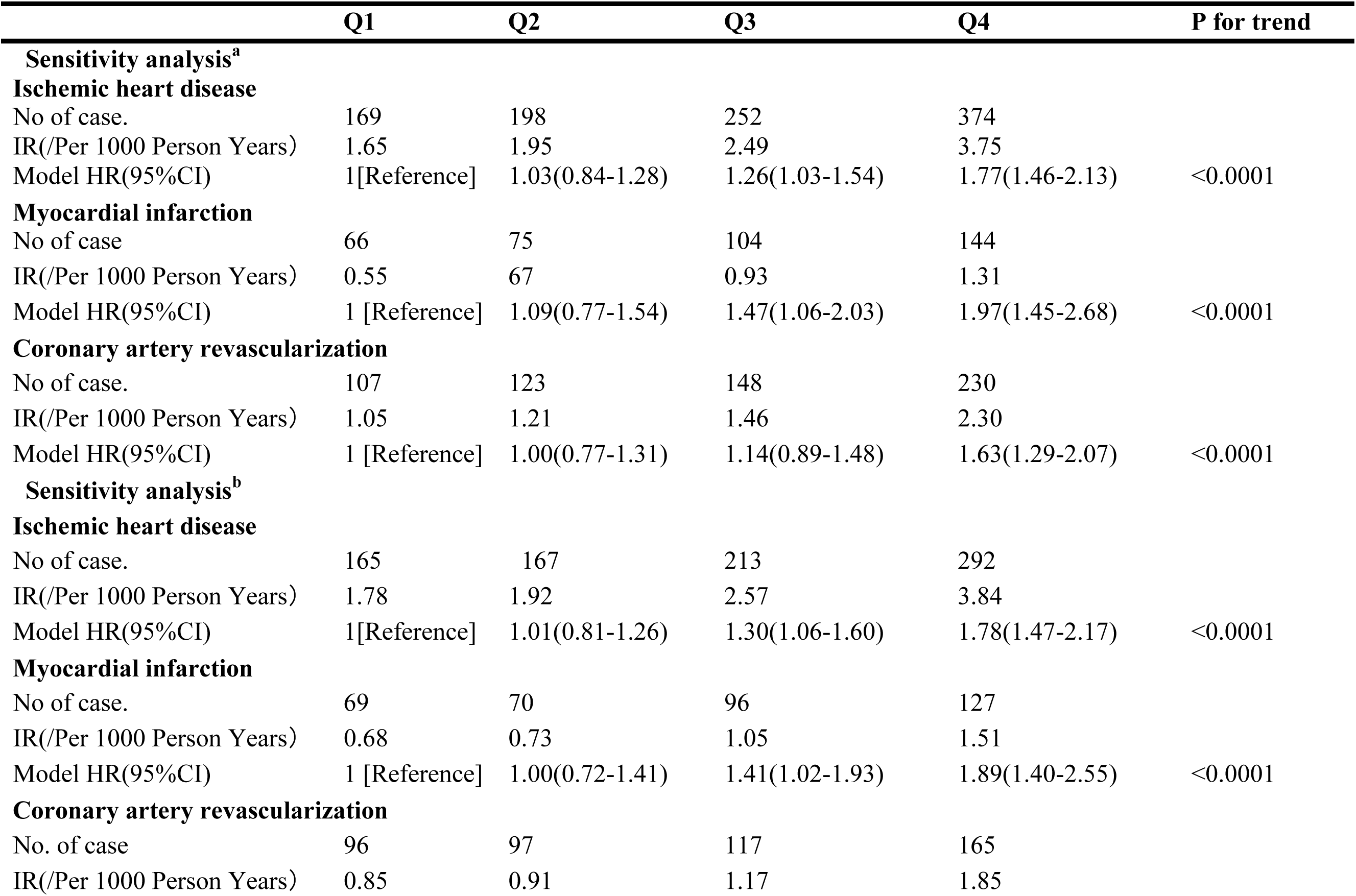
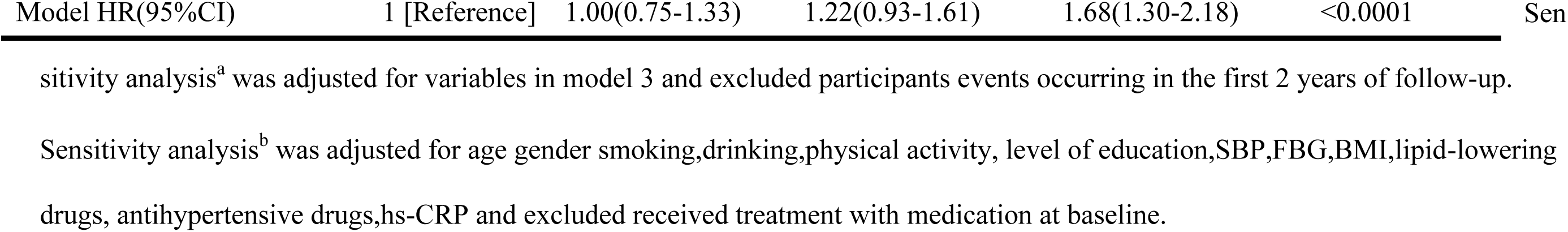
Sensitivity analysis.

**Table 4.** The competing risk of death.

|  | **Q1**<br>**(n=10659)** | **Q2**<br>**(n=10660)** | **Q3**<br>**(n=10660)** | **Q4**<br>**(n=10660)** | **P for trend** |
| --- | --- | --- | --- | --- | --- |
| **Ischemic heart disease** |  |  |  |  |  |
| No of case. | 205 | 245 | 315 | 442 |  |
| IR(/Per 1000 Person Years) | 2.00 | 2.39 | 3.11 | 4.32 |  |
| Model HR(95%CI) | 1 [Reference] | 1.09(0.90-1.31) | 1.33(1.11-1.60) | 1.78(1.50-2.11) | <0.0001 |
| **Myocardial infarction** |  |  |  |  |  |
| No of case. | 81 | 99 | 138 | 181 |  |
| IR(/Per 1000 Person Years) | 0.72 | 0.88 | 1.23 | 1.64 |  |
| Model HR(95%CI) | 1 [Reference] | 1.10(0.92-1.33) | 1.31(1.10-1.57) | 1.87(1.57-2.22) | <0.0001 |
| **Coronary artery revascularization** |  |  |  |  |  |
| No of case. | 124 | 144 | 177 | 261 |  |
| IR(/Per 1000 Person Years) | 1.21 | 1.41 | 1.74 | 2.61 |  |
| Model HR(95%CI) | 1 [Reference] | 1.09(0.91-1.32) | 1.32(1.11-1.58) | 1.80(1.52-2.14) | <0.0001 |

Model was adjusted for covariates in Model 3.

### Stratified analysis

We identified significant interactions between cumRC and factors such as gender, smoking status, and diabetes (P for interaction <0.0001 for all categories). Upon stratifying participants based on gender, smoking habits, and diabetes status, we found that cumRC significantly influenced the risk of IHD in males, especially in the Q4 group (HR, 1.71; 95% CI, 1.42-2.05). However, this correlation was not evident in females. Moreover, the risk of IHD was markedly higher among smokers and individuals with diabetes. Specifically, the HR and 95% CI for the Q4 group were (HR, 1.82; 95% CI, 1.34-2.48) for smokers and (HR, 1.96; 95% CI, 1.34-2.96) for those with diabetes (Table 5).

**Table 5.**
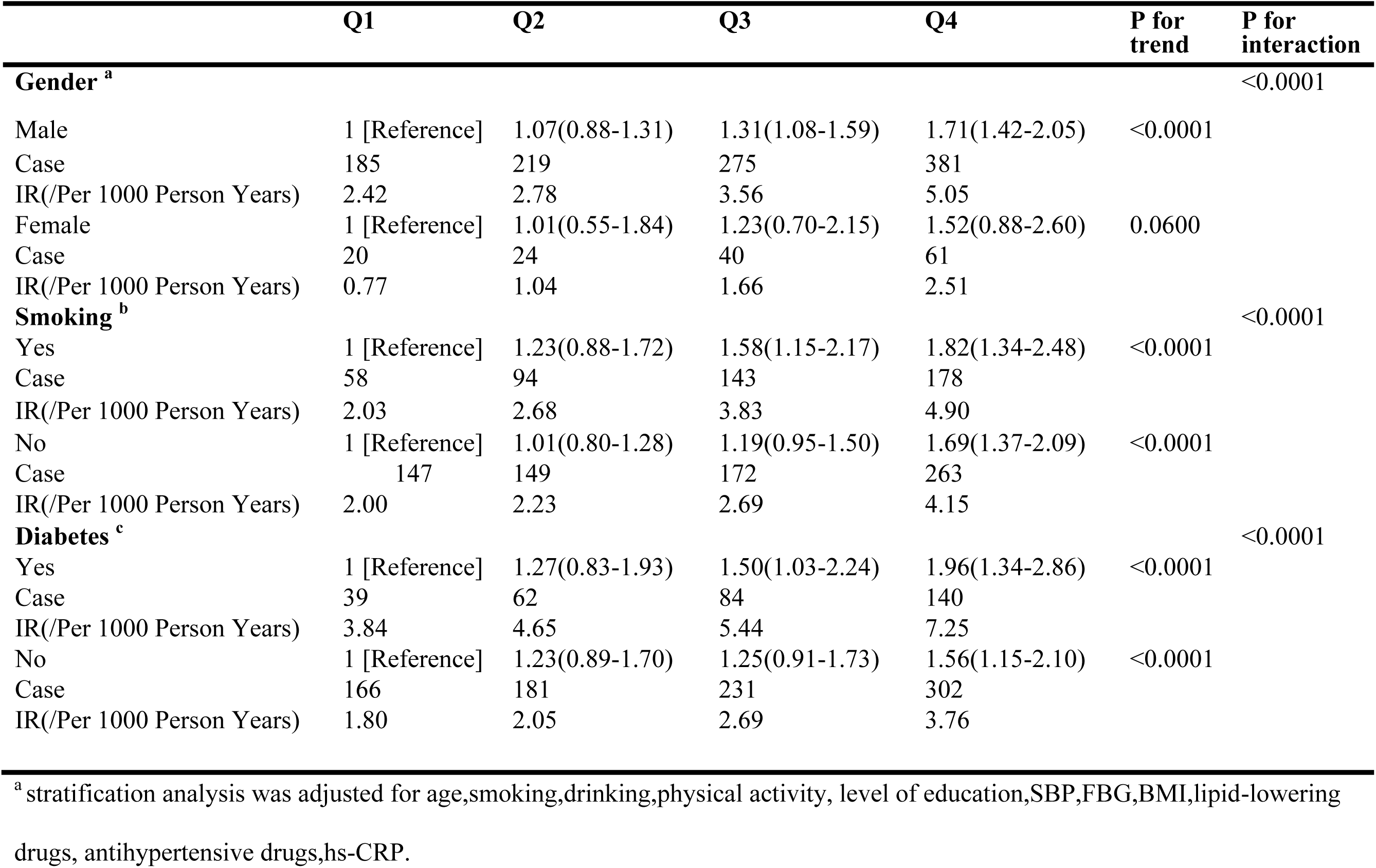
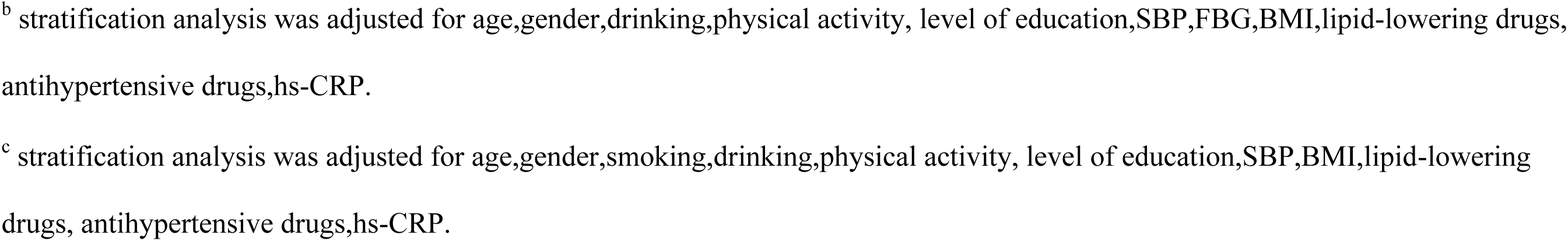
Adjusted HR(95%CI) for incidence of Ischemic heart disease in the all population by gender, smoking status and dibetes.

|  | Q1 | Q2 | Q3 | Q4 | P for trend | P for interaction |
| --- | --- | --- | --- | --- | --- | --- |
| **Gender <sup>a</sup>** |  |  |  |  |  | <0.0001 |
| Male | 1 [Reference] | 1.07(0.88-1.31) | 1.31(1.08-1.59) | 1.71(1.42-2.05) | <0.0001 |  |
| Case | 185 | 219 | 275 | 381 |  |  |
| IR(/Per 1000 Person Years) | 2.42 | 2.78 | 3.56 | 5.05 |  |  |
| Female | 1 [Reference] | 1.01(0.55-1.84) | 1.23(0.70-2.15) | 1.52(0.88-2.60) | 0.0600 |  |
| Case | 20 | 24 | 40 | 61 |  |  |
| IR(/Per 1000 Person Years) | 0.77 | 1.04 | 1.66 | 2.51 |  |  |
| **Smoking <sup>b</sup>** |  |  |  |  |  | <0.0001 |
| Yes | 1 [Reference] | 1.23(0.88-1.72) | 1.58(1.15-2.17) | 1.82(1.34-2.48) | <0.0001 |  |
| Case | 58 | 94 | 143 | 178 |  |  |
| IR(/Per 1000 Person Years) | 2.03 | 2.68 | 3.83 | 4.90 |  |  |
| No | 1 [Reference] | 1.01(0.80-1.28) | 1.19(0.95-1.50) | 1.69(1.37-2.09) | <0.0001 |  |
| Case | 147 | 149 | 172 | 263 |  |  |
| IR(/Per 1000 Person Years) | 2.00 | 2.23 | 2.69 | 4.15 |  |  |
| **Diabetes <sup>c</sup>** |  |  |  |  |  | <0.0001 |
| Yes | 1 [Reference] | 1.27(0.83-1.93) | 1.50(1.03-2.24) | 1.96(1.34-2.86) | <0.0001 |  |
| Case | 39 | 62 | 84 | 140 |  |  |
| IR(/Per 1000 Person Years) | 3.84 | 4.65 | 5.44 | 7.25 |  |  |
| No | 1 [Reference] | 1.23(0.89-1.70) | 1.25(0.91-1.73) | 1.56(1.15-2.10) | <0.0001 |  |
| Case | 166 | 181 | 231 | 302 |  |  |
| IR(/Per 1000 Person Years) | 1.80 | 2.05 | 2.69 | 3.76 |  |  |

<sup>a</sup> stratification analysis was adjusted for age,smoking,drinking,physical activity, level of education,SBP,FBG,BMI,lipid-lowering drugs, antihypertensive drugs,hs-CRP.
<sup>b</sup> stratification analysis was adjusted for age,gender,drinking,physical activity, level of education,SBP,FBG,BMI,lipid-lowering drugs, antihypertensive drugs,hs-CRP.
<sup>c</sup> stratification analysis was adjusted for age,gender,smoking,drinking,physical activity, level of education,SBP,BMI,lipid-lowering drugs, antihypertensive drugs,hs-CRP.

### C-Index, ID and NRI

We incorporated the initial remnant cholesterol level (RC2006), the baseline remnant cholesterol level (RC2010), and cumRC into the baseline model individually. To compare the predictive value of IHD across these three independent factors, we employed the C-index, IDI, and NRI metrics. Our observations revealed that cumRC showcased superior predictive capability for IHD onset, with a C-index of 73.61%. This was in contrast to the foundational model (C-index of 72.85%), the RC2006 prediction model (C-index of 73.57%), and the RC2010 prediction model (C-index of 73.36%). The improvements in the C-index were 0.76%, 0.04%, and 0.25% respectively. Furthermore, the IDI values for all three models exceeded 0. When juxtaposed with the foundational model, the NRI values witnessed an increment of 14.10%, 20.25%, and 20.31% respectively (all P < 0.0001). (Refer to Table 6).

**Table 6.** use C-index、IDI、NRI Compare the risk prediction value of incidence Ischemic heart disease.

|  | **C-index** | **IDI** | **P for IDI** | **NRI** | **P for NRI** |
| --- | --- | --- | --- | --- | --- |
| Model | 0.7285 | 1(reference) |  | 1(reference) |  |
| Model+RC<sub>2006</sub> | 0.7357 | 0.000735 | 0.0002 | 0.1410 | <0.0001 |
| Model+RC<sub>2010</sub> | 0.7336 | 0.000787 | <0.0001 | 0.2025 | <0.0001 |
| Model+cumRC | 0.7361 | 0.001029 | <0.0001 | 0.2031 | <0.0001 |

RC<sub>2006</sub>,the initial remnant cholesterol level.RC<sub>2010</sub>,the baseline remnant cholesterol level.
Model,adjusted for variables in model 3.

## DISCUSSION

In our comprehensive prospective cohort study, we discerned a strong correlation between elevated cumRC and an augmented risk of emerging IHD. Notably, this relationship persisted even after accounting for LDL-C and HDL-C levels. When juxtaposed with a singular RC measurement, cumRC emerged as a superior predictor for IHD onset. Moreover, the risk of IHD manifestation was notably higher among smokers and individuals with diabetes.

Our study indicates that higher cumRC levels correlate with an increased likelihood of IHD incidents, and this correlation remains consistent even when considering traditional risk factors. When compared to the reference group with the lowest cumRC levels (Q1 group), the group with the highest cumRC levels (Q4 group) showed a 69% surge in risk. Moreover, with the rise in cumRC levels, the IHD risk also exhibited an upward trajectory (P < 0.0001), hinting at a dose-response relationship between cumRC and IHD risk. While the link between cumRC and IHD hasn’t been the focal point of past research, there’s evidence suggesting that a single-measurement of elevated RC is tied to a heightened IHD risk. For instance, Joshi, Parag H et al. [23], in their analysis of 818 participants from the Framingham Offspring Cohort Study and 4114 from the Jackson Heart Study, found that a 1-SD increase in RC corresponded to a 23% uptick in coronary heart disease risk. In a broader context, a study from Copenhagen revealed that individuals exposed to high RC levels faced a steeper risk of myocardial infarction; specifically, compared to RC levels < 0.5 mmol/L, the risk amplified 4.2 times when RC reached or exceeded 1.5 mmol/L.[24] Another study posits that when RC levels surpass the median (<0.6 mmol/L), the IHD risk for those in the 75th percentile (1.0 mmol/L) and 95th percentile (1.6 mmol/L) jumps by 43% and 49%, respectively.[25] These insights align with and bolster our findings. Yet, it’s worth noting that most prior studies zoomed in on single-measurement RC levels, overlooking the prolonged influence of cumRC on IHD. Our research bridges this gap and underscores the importance of not just regular RC monitoring but also heightened scrutiny of cumRC to pinpoint individuals with a pronounced IHD risk.

Furthermore, our research highlights a pronounced association between cumRC and IHD risk, especially among smokers and individuals with diabetes. Earlier studies have shown that smokers face a 50% higher risk of IHD compared to non-smokers.[26] A study from Japan further emphasizes this, indicating that among those with elevated cholesterol, smokers are 2.68 times more likely to develop coronary heart disease than non-smokers.[27] In our findings, when compared to the group with low cumRC levels, the risk of IHD surged by 82% among smokers and 69% among non-smokers.

In the context of diabetes, our study indicates that individuals in the highest cumRC quartile face a risk of IHD that’s almost twice (1.96-fold) that of those in the lowest quartile. For those without diabetes, the risk is increased by 1.56-fold. Another study has shown that, relative to a serum cholesterol baseline of <5.2 mmol/L, having levels >7.2 mmol/L elevates the risk of coronary heart disease mortality by 78% in diabetics and 23% in non-diabetics.[28] A cohort study from South Korea further supports this, revealing that among type 2 diabetics, those in the highest RC quartile have a 1.281-fold increased risk of cardiovascular disease compared to the lowest quartile.[29]These findings underscore the urgency of rigorous lipid management, especially for high-risk groups like smokers and diabetics, who are already susceptible to atherosclerotic cardiovascular disease (ASCVD).

Previous research indicates that the prolonged cumulative exposure to certain risk factors has a more pronounced impact on adverse outcomes, such as cardiovascular disease, than isolated, one-time exposures. For example, a cohort study with 11,502 participants over an average of 12.9 years revealed that high cumSBP is a significant risk factor for cardiovascular disease. As cumSBP levels rise, so does the risk of cardiovascular disease, even when accounting for individual one-time systolic blood pressure readings. [30]Yiyi Zhang et al. [16] found that increased cumLDL-C exposure heightens the risk of coronary heart disease. When comparing the highest quartile group to the lowest, there was a 1.57-fold increase in the risk. Notably, this association remained strong even after adjusting for individual one-time cumLDL-C measurements, mirroring our study’s findings.Our research determined that higher cumRC levels correlate with an increased IHD risk. When comparing the highest cumRC quartile group to the lowest, the risk of IHD was notably higher (HR, 1.69; 95% CI, 1.42-2.01). This risk remained significant even after adjusting for baseline RC levels (HR, 1.42; 95% CI, 1.18-11.72), suggesting that the influence of cumRC on IHD is independent of individual one-time RC measurements.

Moreover, we developed a risk prediction model using traditional risk factors and incorporated RC measurements from 2006, 2010, and cumRC. The results showed that the cumRC model had a superior predictive capability compared to the individual RC models from 2006 and 2010. This implies that cumRC might offer a more robust predictive power for IHD risk.

We also enhanced the robustness of our findings by conducting a sensitivity analysis. We excluded participants who either developed IHD within the first two years of the study or were on medication. Consequently, when identifying high-risk groups for ASCVD, it’s essential to consider both individual RC and cumRC levels.

While the precise mechanisms linking cumRC to IHD remain unclear, existing research offers several potential explanations for this association. First, similar to LDL-C, RC has been identified as a potential contributor to atherosclerosis.[31, 32] Unlike LDL-C, RC accumulates within vascular walls when present in plasma. It remains unaltered and is absorbed by macrophages and smooth muscle cells, resulting in foam cell formation and subsequent atherosclerosis development.[33] Additionally, RC has up to 40 times more cholesterol than LDL-C, which might make it more susceptible to atherosclerosis initiation.[34]

Second, high RC levels are seen as a potential risk for endothelial dysfunction, which can further advance atherosclerosis.[35] Atherosclerosis is a primary pathological factor in IHD development.[36] However, for vascular lesions to occur, there needs to be an extended exposure to elevated RC levels. A single measurement is insufficient to gauge the damage from prolonged exposure to high RC levels. The progression of IHD might be closely tied to the cumulative exposure to RC. Our study addressed the limitations of single RC measurements by using cumRC, derived from multiple RC readings, to evaluate the IHD risk.

It’s also noteworthy that, even after accounting for hs-CRP levels, our research determined that cumRC remains an independent IHD risk factor. Previous studies have shown that RC can cause mild inflammation and activate endothelial cells, leading to atherosclerosis and subsequently increasing IHD risk.[37]

This study offers multiple advantages. First and foremost, it stands as the inaugural large-scale prospective cohort study delving into the association between cumRC and IHD risk. The substantial sample size and extended follow-up duration further bolster its credibility. Second, our meticulous adjustment for potential confounders, combined with stratified and sensitivity analyses, fortifies the validity of our findings. Third, by collecting multiple RC measurements from participants, our study provides a more dependable insight compared to prior studies that relied on a single measurement. Lastly, the accuracy of our outcome data is ensured by sourcing it from electronic medical records and the social security information system, eliminating the inconsistencies that might arise from participant recollections.

## CONCLUSION

Elevated cumRC levels correlate with a heightened risk of IHD onset. This revelation accentuates the pivotal role of cumRC in preemptive IHD prevention measures. As part of lipid management strategies, prioritizing the surveillance of cumRC is imperative to curtail the IHD risks linked with increased cumRC concentrations.

## DECLARATIONS

### Ethics approval and consent to participate

The project protocol was approved by the ethics committee of Ethics Committee of the Kailuan Medical Group and was by the guidelines of the Helsinki Declaration, and all study individuals in this project signed an informed consent form at enrollment.

### Consent for publication

If the manuscript is accepted, we approve it for publication in Journal of the American Heart Association.

### Availability of data and materials

The data that support the findings of this study are available from [third party name] but restrictions apply to the availability of these data, which were used under license for the current study, and so are not publicly available. Data are however available from the corresponding author upon reasonable request and with permission of the corresponding author.

### Authors’ contributions

XZ made substantial contributions to study design, data collection, data analysis and manuscript writing. SW and XL made substantial contributions to study design and intellectual direction. YW,WL,HG,HW,JY and GW made contributions to data collection. All authors read and approved the final manuscript.

## Non-standard Abbreviations and Acronyms

RC: remnant cholesterol
IHD: ischemic heart disease
cumRC: cumulative remnant cholesterol
LDL-C: lipoprotein cholesterol
ASCVD: atherosclerotic cardiovascular incidents
FBG: Fasting Blood Glucose
TG: Triglycerides
TC: Total Cholesterol
HDL-C: High-Density Lipoprotein Cholesterol
SD: standard deviation
BMI: Body Mass Index
DBP: Diastolic Blood Pressure
HR: Hazard Ratio

## Acknowledgements

The authors thank the investigators who made this cohort study possible.

## REFERENCES

1. Valdes-Marquez E, Parish S, Clarke R, Stari T, Worrall BB, ISGC MCot, et al. Relative effects of ldl-c on ischemic stroke and coronary disease: A mendelian randomization study. Neurology. 2019;92:e1176–e1187

2. Grundy SM, Stone NJ, Bailey AL, Beam C, Birtcher KK, Blumenthal RS, et al. 2018 aha/acc/aacvpr/aapa/abc/acpm/ada/ags/apha/aspc/nla/pcna guideline on the management of blood cholesterol: A report of the american college of cardiology/american heart association task force on clinical practice guidelines. Circulation. 2019;139:e1082–e1143

3. Ference BA, Ginsberg HN, Graham I, Ray KK, Packard CJ, Bruckert E, et al. Low-density lipoproteins cause atherosclerotic cardiovascular disease. 1. Evidence from genetic, epidemiologic, and clinical studies. A consensus statement from the european atherosclerosis society consensus panel. Eur Heart J. 2017;38:2459–2472

4. Schubert J, Lindahl B, Melhus H, Renlund H, Leosdottir M, Yari A, et al. Low-density lipoprotein cholesterol reduction and statin intensity in myocardial infarction patients and major adverse outcomes: A swedish nationwide cohort study. Eur Heart J. 2021;42:243–252

5. Sandesara PB, Virani SS, Fazio S, Shapiro MD. The forgotten lipids: Triglycerides, remnant cholesterol, and atherosclerotic cardiovascular disease risk. Endocr Rev. 2019;40:537–557

6. Varbo A, Nordestgaard BG. Remnant lipoproteins. Curr Opin Lipidol. 2017;28:300–307

7. Twickler TB, Dallinga-Thie GM, Cohn JS, Chapman MJ. Elevated remnant-like particle cholesterol concentration: A characteristic feature of the atherogenic lipoprotein phenotype. Circulation. 2004;109:1918–1925

8. Varbo A, Benn M, Tybjaerg-Hansen A, Jorgensen AB, Frikke-Schmidt R, Nordestgaard BG. Remnant cholesterol as a causal risk factor for ischemic heart disease. J Am Coll Cardiol. 2013;61:427–436

9. Castaner O, Pinto X, Subirana I, Amor AJ, Ros E, Hernaez A, et al. Remnant cholesterol, not ldl cholesterol, is associated with incident cardiovascular disease. J Am Coll Cardiol. 2020;76:2712–2724

10. Gao S, Xu H, Ma W, Yuan J, Yu M. Remnant cholesterol predicts risk of cardiovascular events in patients with myocardial infarction with nonobstructive coronary arteries. J Am Heart Assoc. 2022;11:e024366

11. Kaltoft M, Langsted A, Nordestgaard BG. Triglycerides and remnant cholesterol associated with risk of aortic valve stenosis: Mendelian randomization in the copenhagen general population study. Eur Heart J. 2020;41:2288–2299

12. Yu D, Wang Z, Zhang X, Qu B, Cai Y, Ma S, et al. Remnant cholesterol and cardiovascular mortality in patients with type 2 diabetes and incident diabetic nephropathy. J Clin Endocrinol Metab. 2021;106:3546–3554

13. Quispe R, Martin SS, Michos ED, Lamba I, Blumenthal RS, Saeed A, et al. Remnant cholesterol predicts cardiovascular disease beyond ldl and apob: A primary prevention study. Eur Heart J. 2021;42:4324–4332

14. Navar-Boggan AM, Peterson ED, D’Agostino RB, Sr., Neely B, Sniderman AD, Pencina MJ. Hyperlipidemia in early adulthood increases long-term risk of coronary heart disease. Circulation. 2015;131:451–458

15. Domanski MJ, Tian X, Wu CO, Reis JP, Dey AK, Gu Y, et al. Time course of ldl cholesterol exposure and cardiovascular disease event risk. J Am Coll Cardiol. 2020;76:1507–1516

16. Zhang Y, Pletcher MJ, Vittinghoff E, Clemons AM, Jacobs DR, Jr., Allen NB, et al. Association between cumulative low-density lipoprotein cholesterol exposure during young adulthood and middle age and risk of cardiovascular events. JAMA Cardiol. 2021;6:1406–1413

17. Wu Z, Jin C, Vaidya A, Jin W, Huang Z, Wu S, et al. Longitudinal patterns of blood pressure, incident cardiovascular events, and all-cause mortality in normotensive diabetic people. Hypertension. 2016;68:71–77

18. Wu S, Huang Z, Yang X, Zhou Y, Wang A, Chen L, et al. Prevalence of ideal cardiovascular health and its relationship with the 4-year cardiovascular events in a northern chinese industrial city. Circ Cardiovasc Qual Outcomes. 2012;5:487–493

19. Polonikov AV, Vialykh EK, Churnosov MI, Illig T, Freidin MB, Vasil’eva OV, et al. The c718t polymorphism in the 3’- untranslated region of glutathione peroxidase-4 gene is a predictor of cerebral stroke in patients with essential hypertension. Hypertens Res. 2012;35:507–512

20. Genuth S, Alberti KG, Bennett P, Buse J, Defronzo R, Kahn R, et al. Follow-up report on the diagnosis of diabetes mellitus. Diabetes care. 2003;26:3160-3167

21. Pool LR, Ning H, Wilkins J, Lloyd-Jones DM, Allen NB. Use of long-term cumulative blood pressure in cardiovascular risk prediction models. JAMA Cardiol. 2018;3:1096–1100

22. Tunstall-Pedoe H, Kuulasmaa K, Amouyel P, Arveiler D, Rajakangas AM, Pajak A. Myocardial infarction and coronary deaths in the world health organization monica project. Registration procedures, event rates, and case-fatality rates in 38 populations from 21 countries in four continents. Circulation. 1994;90:583–612

23. Joshi PH, Khokhar AA, Massaro JM, Lirette ST, Griswold ME, Martin SS, et al. Remnant lipoprotein cholesterol and incident coronary heart disease: The jackson heart and framingham offspring cohort studies. Journal of the American Heart Association. 2016;5

24. Wadstrom BN, Wulff AB, Pedersen KM, Jensen GB, Nordestgaard BG. Elevated remnant cholesterol increases the risk of peripheral artery disease, myocardial infarction, and ischaemic stroke: A cohort-based study. Eur Heart J. 2022;43:3258–3269

25. Doi T, Langsted A, Nordestgaard BG. Elevated remnant cholesterol reclassifies risk of ischemic heart disease and myocardial infarction. J Am Coll Cardiol. 2022;79:2383–2397

26. Jee Y, Jung KJ, Lee S, Back JH, Jee SH, Cho SI. Smoking and atherosclerotic cardiovascular disease risk in young men: The korean life course health study. BMJ Open. 2019;9:e024453

27. Hata J, Doi Y, Ninomiya T, Fukuhara M, Ikeda F, Mukai N, et al. Combined effects of smoking and hypercholesterolemia on the risk of stroke and coronary heart disease in japanese: The hisayama study. Cerebrovasc Dis. 2011;31:477–484

28. Adlerberth AM, Rosengren A, Wilhelmsen L. Diabetes and long-term risk of mortality from coronary and other causes in middle-aged swedish men. A general population study. Diabetes care. 1998;21:539–545

29. Huh JH, Han KD, Cho YK, Roh E, Kang JG, Lee SJ, et al. Remnant cholesterol and the risk of cardiovascular disease in type 2 diabetes: A nationwide longitudinal cohort study. Cardiovasc Diabetol. 2022;21:228

30. Reges O, Ning H, Wilkins JT, Wu CO, Tian X, Domanski MJ, et al. Association of cumulative systolic blood pressure with long-term risk of cardiovascular disease and healthy longevity. Hypertension. 2021;77:347–356

31. Ference BA, Kastelein JJP, Ray KK, Ginsberg HN, Chapman MJ, Packard CJ, et al. Association of triglyceride-lowering lpl variants and ldl-c-lowering ldlr variants with risk of coronary heart disease. JAMA. 2019;321:364–373

32. Chapman MJ, Ginsberg HN, Amarenco P, Andreotti F, Borén J, Catapano AL, et al. Triglyceride-rich lipoproteins and high-density lipoprotein cholesterol in patients at high risk of cardiovascular disease: Evidence and guidance for management. European Heart Journal. 2011;32:1345–1361

33. Nakajima K, Nakano T, Tanaka A. The oxidative modification hypothesis of atherosclerosis: The comparison of atherogenic effects on oxidized ldl and remnant lipoproteins in plasma. Clin Chim Acta. 2006;367:36–47

34. Boren J, Williams KJ. The central role of arterial retention of cholesterol-rich apolipoprotein-b-containing lipoproteins in the pathogenesis of atherosclerosis: A triumph of simplicity. Curr Opin Lipidol. 2016;27:473–483

35. Nakamura T, Takano H, Umetani K, Kawabata K, Obata JE, Kitta Y, et al. Remnant lipoproteinemia is a risk factor for endothelial vasomotor dysfunction and coronary artery disease in metabolic syndrome. Atherosclerosis. 2005;181:321–327

36. Konishi M, Iso H, Komachi Y, Iida M, Shimamoto T, Jacobs DR, Jr., et al. Associations of serum total cholesterol, different types of stroke, and stenosis distribution of cerebral arteries. The akita pathology study. Stroke. 1993;24:954–964

37. Varbo A, Benn M, Tybjaerg-Hansen A, Nordestgaard BG. Elevated remnant cholesterol causes both low-grade inflammation and ischemic heart disease, whereas elevated low-density lipoprotein cholesterol causes ischemic heart disease without inflammation. Circulation. 2013;128:1298–1309

38. Varbo A, Nordestgaard BG. Directly measured vs. Calculated remnant cholesterol identifies additional overlooked individuals in the general population at higher risk of myocardial infarction. Eur Heart J. 2021;42:4833–4843

